# South Asia’s unprotected poor: a systematic review of why social protection programs fail to reach their potential

**DOI:** 10.1101/2023.11.23.23298962

**Authors:** Warda Javed, Zubia Mumtaz

**Affiliations:** School of Public Health, University of Alberta, 3-309 Edmonton Clinic Health Academy, 11405 – 87 Ave, Edmonton, AB T6G 1C9, Canada

## Abstract

The incongruity between South Asia’s economic growth and extreme poverty has led to a growing interest in social protection and the subsequent implementation of anti-poverty programs. These work to promote inclusive growth and ensure that the poor do not get left behind. However, many programs have systematically failed to achieve their full potential in reaching the poorest of the poor. We reviewed the literature to understand the determinants behind this inequity in South Asia.

A search of four databases, EconLit, Global Health Database, MEDLINE, SocINDEX, supplemented by citation tracking and an external search, yielded 42 papers evaluating 23 social protection programs. All articles were assessed for quality using the GRADE and GRADE CERQual criteria. Data were analyzed using Thomas & Harding’s thematic synthesis approach to generate new higher-order interpretations.

Our analysis identified five themes underscoring program processes that stop resources from reaching the poor. These include: (1) structurally flawed program theories that overlook the complexities of poverty and are instead rooted in simplistic cause-and-effect approaches overestimating the poor’s gain from participation; (2) elite capture of program resources through the direct appropriation of benefits, their powerful positioning in program implementation, and their ability to dictate the poor’s accessibility through relationships of patronage and withholding of information; (3) insufficient targeting strategies to reach the poorest and a subsequent redirection of resources toward the rich; (4) program designs that overlook gender-based restrictions, hidden costs, the poor’s lack of legal documentation, and their physical and social exclusion; (5) some of the poorest households actively choosing self-exclusion from social protection due to a desire to maintain dignity, a lack of capital, and a perception of programs as substandard.

The review highlights the disconnect between social protection program designs and the ground realities of their ‘ideal’ beneficiaries: the poorest of the poor. We propose the persistence of this well-documented disconnect may stem from three sources. First, there is an unclear understanding of who the poor are in South Asia, with definitions overlooking the historical influence of the caste system. Western perceptions of poverty continue to dominate the discourse. The second challenge is effective engagement and co-production of knowledge with the poor. Lastly, despite encouragement of international collaboration, fast-paced funding calls do not allocate sufficient time to build relationships with the poor primary stakeholders. We suggest the possibility that maintenance of this disconnect is intentional, reflecting a broader power dynamic in which the global and local elite dictate the lives of the poor based on geopolitical interests and national priorities.

## Introduction

Over the past few decades, substantial economic growth has been achieved globally, most notably in low and middle-income countries [1,2]. Before the COVID-19 pandemic, the growth was largely accompanied by a concomitant decline in poverty trends [3]. That said, poverty reduction was lower than expected, considering the rate of fiscal expansion. A failure of the trickle-down theory, a lack of pro-poor investment in the social sector, and globalization further left the poor behind and amplified the inequities [1,2,4,5].

This failure to reduce poverty led to a growing interest in social protection, a broad term denoting programs and policies that “[tackle] poverty and vulnerability while strengthening inclusive social development and equitable economic growth.” [6] Usually an *ex-post* response to poverty, the programs intend to establish inclusive growth via promotive, protective [7], preventive [8], and transformative anti-poverty interventions [9]. They include the instruments of social assistance/social safety nets, social care, social insurance, and labour market policies [10]. Through short-term responses to shocks and long-term sustainable and systemic interventions, these strategies aim to support an individual’s movement out of poverty or prevent it altogether [11].

The demand for social protection has heightened due to the multiple overlapping crises of climate change, deglobalization, and, most recently, COVID-19. The pandemic alone has added 124 million individuals [12] to the 648 million [3] already living in extreme poverty, particularly in the world’s resource-poor regions. Social assistance has become the primary tool to cushion these impacts, forming 78% of state-implemented COVID-19 relief [13]. According to recent data, over 1,700 new social protection initiatives were introduced to address the pandemic alone [12].

However, emerging evidence suggests social protection programs fail to realize their potential in targeting the poorest of the poor. A 2022 United Nations report [14] urges that “little attention has been paid to the countless individuals that are falling through the cracks of [social protection].” Kidd & Athias [15] echo these apprehensions in their evaluation of 38 programs across 23 low- and middle-income countries. They show that targeted initiatives did not reach over half the poorest quintile of their anticipated beneficiaries. The Atlas of Social Protection Indicators of Resilience and Equity dataset from 2014-2019 paralleled these findings, showing that social assistance programs in low-income countries covered only 17% of the poorest households [16].

It is particularly important to focus on South Asia, a region home to 21% of the world’s poor [3]. It is plagued with high national debts and inflation [17], corruption, and conflict [18]. South Asia is also on the frontline of the impacts of climate change [19]. Despite the economic progress of some countries, notably India and Bangladesh [20], it remains a region characterized by deeply rooted systemic inequities that subordinate and marginalize some subgroups [21,22,23]. This marginalization is structured within historical caste-based social hierarchies, which are reinforced by a tangible reality in which the poor have limited rights to engage in the economic, social, and political spheres of their society [24]. An example of this exclusion is the failure of Pakistan’s Benazir Income Support Programme and India’s Mahatma Gandhi National Rural Employment Guarantee Scheme to reach, respectively, 79% and 68% of the poorest quintile of their target populations [15]. Only 25% of Pakistan’s poorest households participate in a social protection program [25].

Social protection is a potentially invaluable tool to address poverty and vulnerability within a world undergoing major changes - whether that be to its climate, AI technologies, or the post-pandemic upheaval. Given the evidence that deserving individuals do not receive their entitled benefits, we reviewed the literature to explore why programs do not reach the poorest of the poor and realize their full potential. As social protection is a vast domain, our review will focus on non-contributory social assistance (e.g., cash transfers, food subsidies, public works programs), microfinance programs, and social health protection. Each of these branches falls within our interests of state-sanctioned and long-term resources targeted toward the poorest of the poor.

## Methods

A formal search of four databases - EconLit, Global Health Database, MEDLINE, SocINDEX - yielded 497 articles. See Table 1 for a list of search terms. These papers were first filtered by title and abstract; 154 papers were assessed for eligibility based on full-text review and 30 met our inclusion criteria. See Table 2 for inclusion and exclusion criteria. We supplemented the search through citation tracking from reference sections of our selected papers. Additional supplementary searches about specific programs were performed as needed using Google. This yielded an additional 12 papers. Figure 1 presents the Preferred Reporting Items for Systematic Reviews and Meta-analysis (PRISMA) [26] flow diagram detailing our search process. Ethics clearance was received from the University of Alberta’s Alberta Research Information Services (ARISE) System (PRO#00117838).

**Figure 1.** PRISMA 2020 Flow Diagram.

**Table 1.**
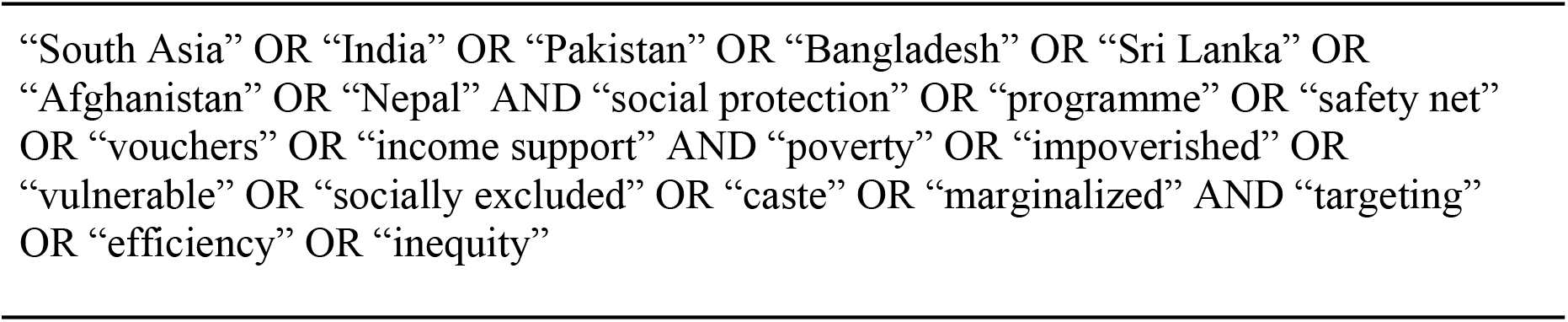
Keywords in Literature Search.

**Table 2.** Inclusion and Exclusion Criteria for Systematic Review.

| Inclusion Criteria | Exclusion Criteria |
| --- | --- |
| Published in the peer-reviewed literature only. | Non-English language. |
| English language between 2000-2023. | Non-peer-reviewed or non-empirical publications (i.e., evaluation reports, working papers, reviews, commentaries, news items, magazines). |
| Qualitatively or quantitatively evaluates factors that inhibit South Asian poor from accessing social protection programs. | Theoretical and econometric-driven research. |
| Acknowledges and highlights the role of social and political factors in program outcomes. | Short-term programs implemented by non-governmental or civil society organizations. |
|  | Publications not evaluating South Asian social protection programs targeted toward the poor. |
|  | Evaluation of program outputs only, failing to assess processes related to targeting the poor. |

Descriptive data were extracted from each study, including the country of the social protection program, research method, sample size, and key findings. The Grading of Recommendations, Assessment, Development, and Evaluations (GRADE) [27] and GRADE-CERQual (Confidence in the Evidence from Reviews of Qualitative Research) [28] criteria were used to assess the quality of quantitative and qualitative studies, respectively. Mixed-method studies were evaluated using both criteria. Following the appraisal, papers were categorized based on their quality score of high, medium, or low. Only high and medium quality score papers were included in the review. See Table 3 for detailed summaries of all papers.

**Table 3.**
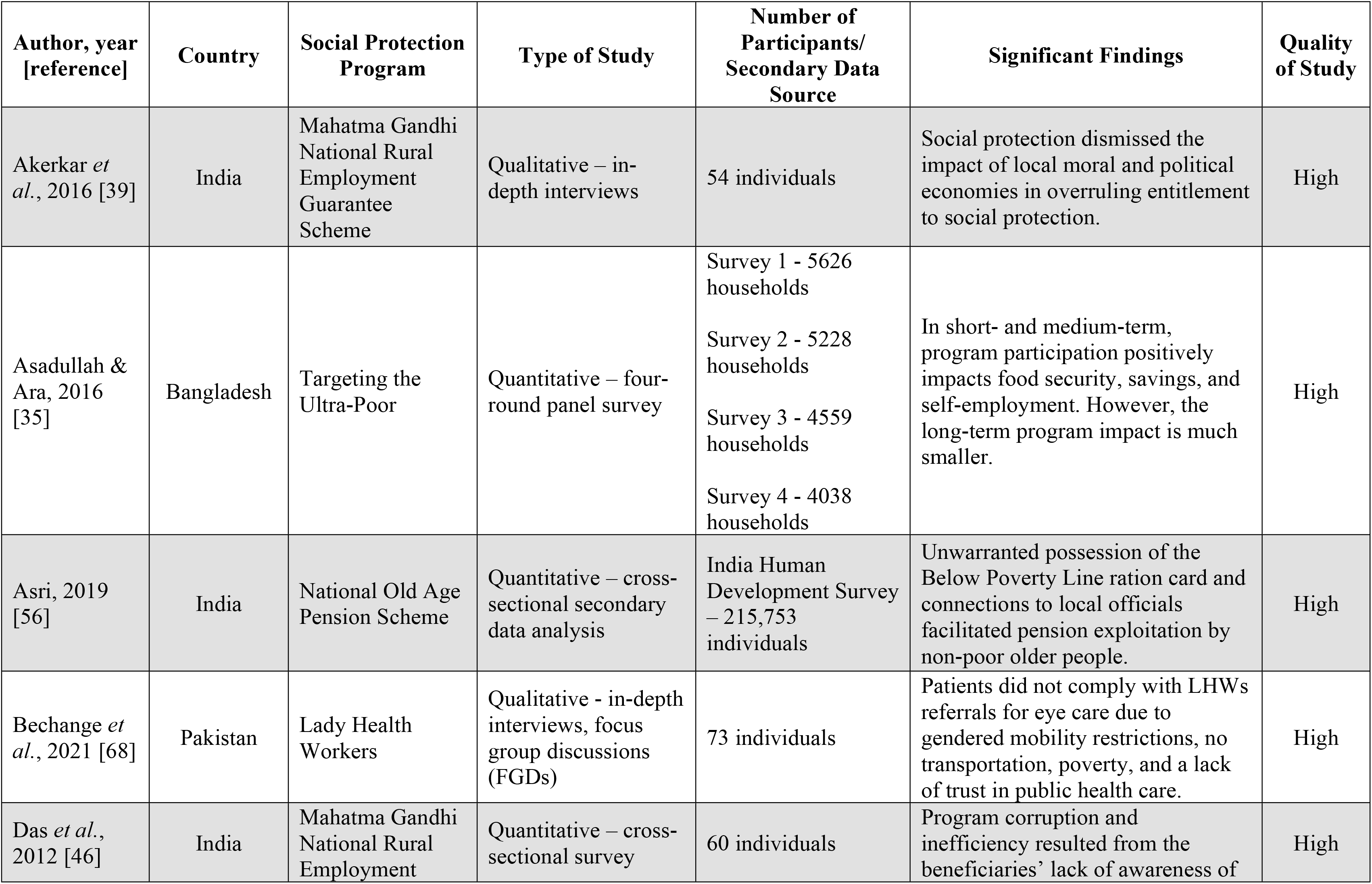
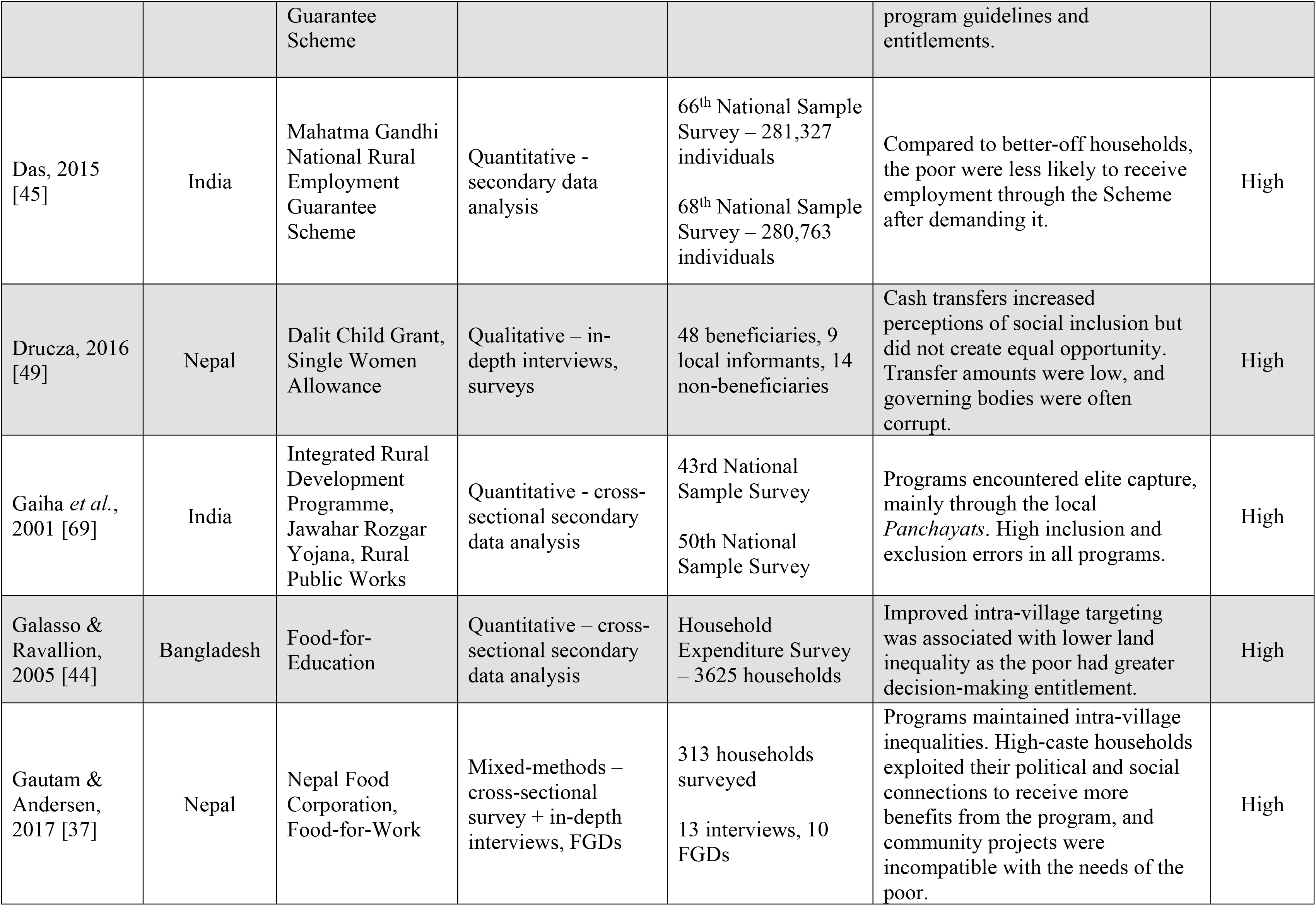
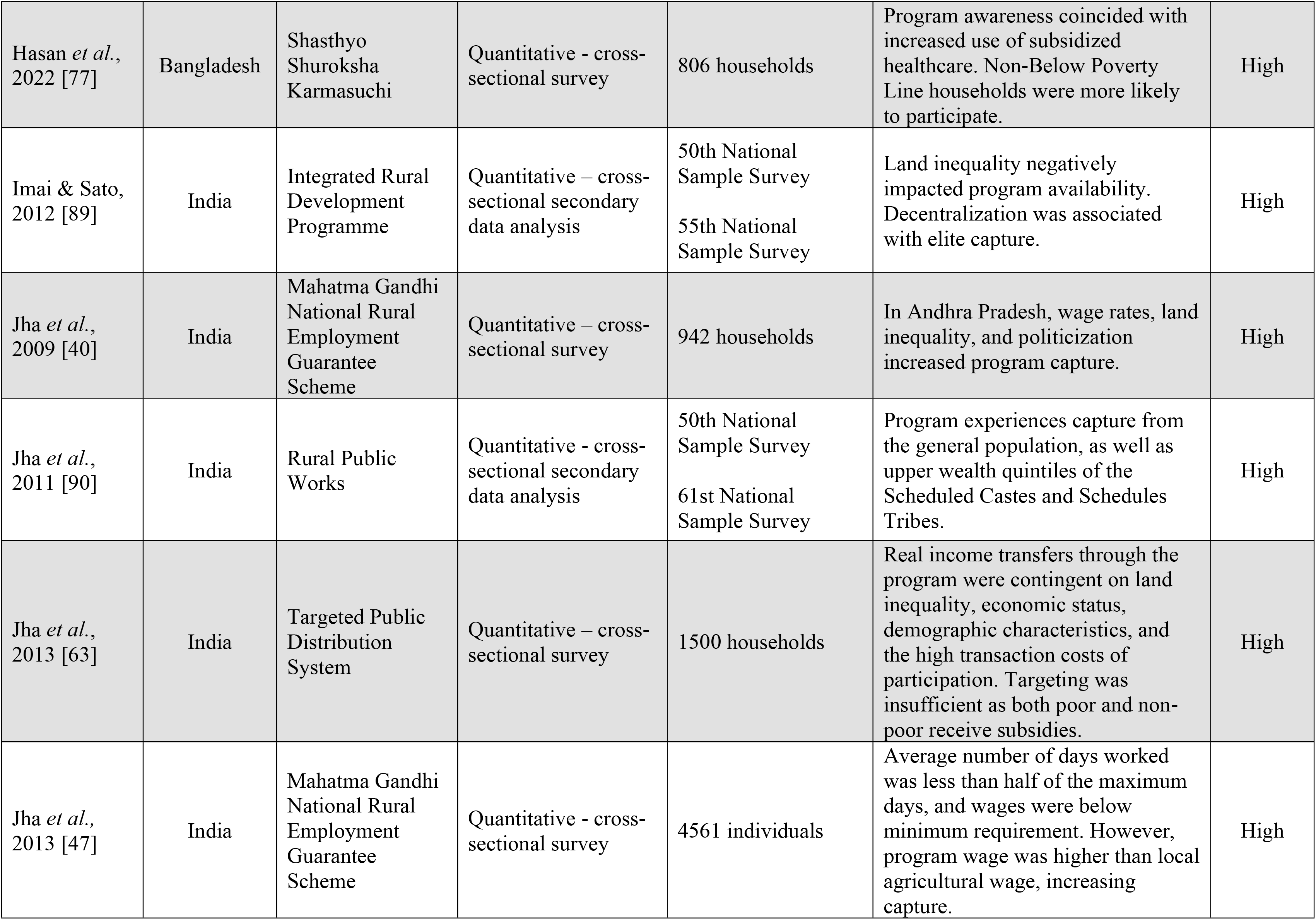
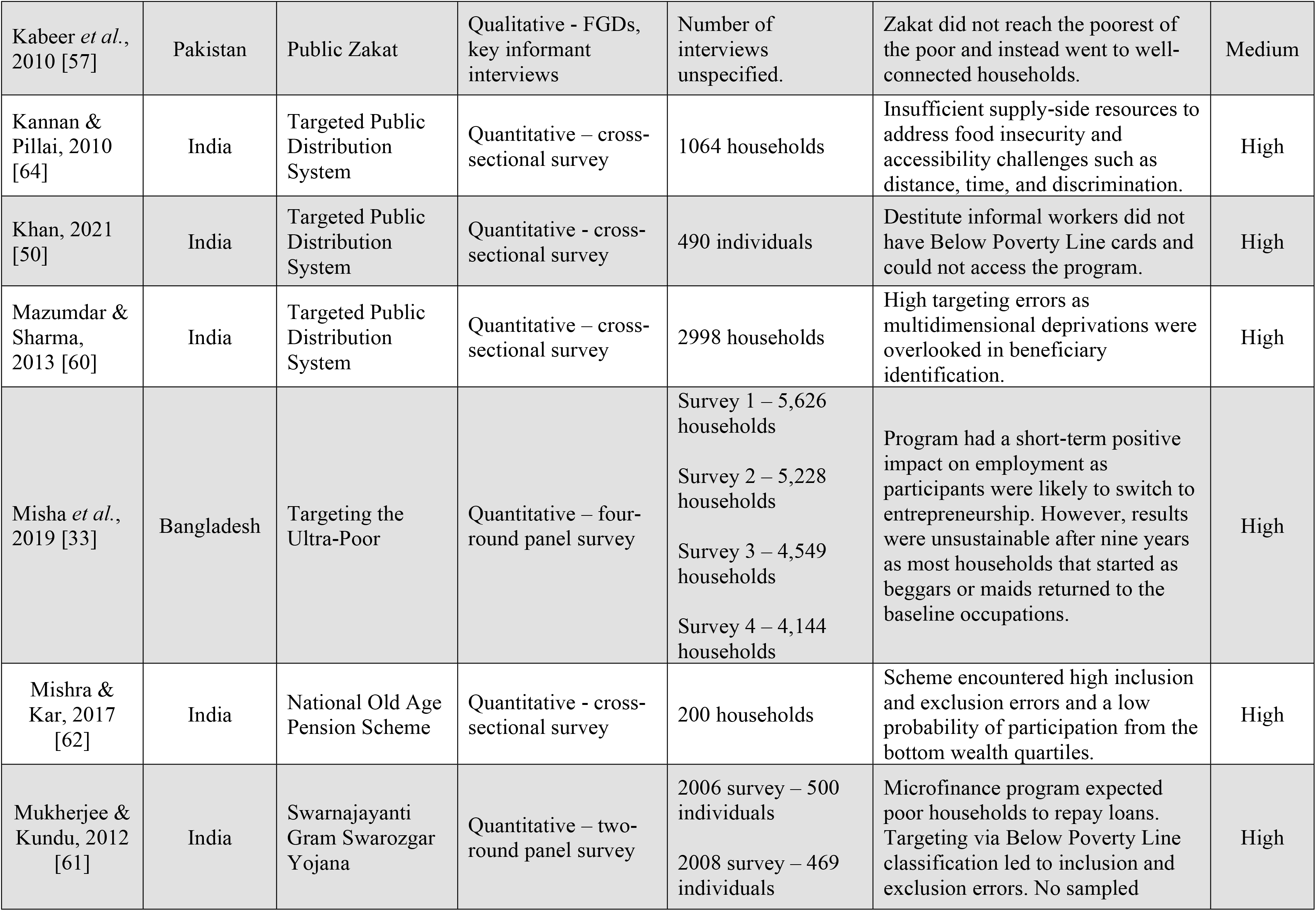
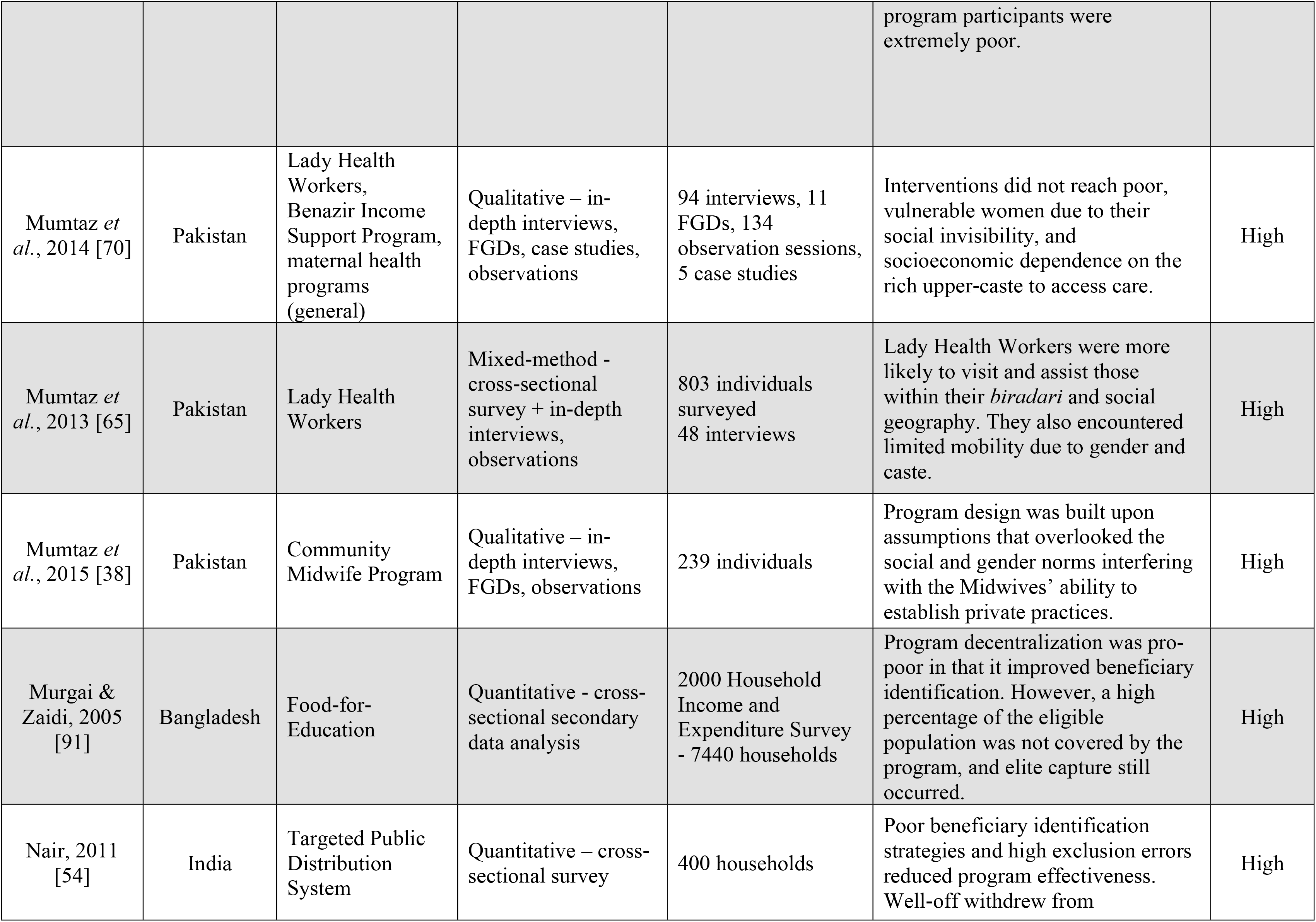
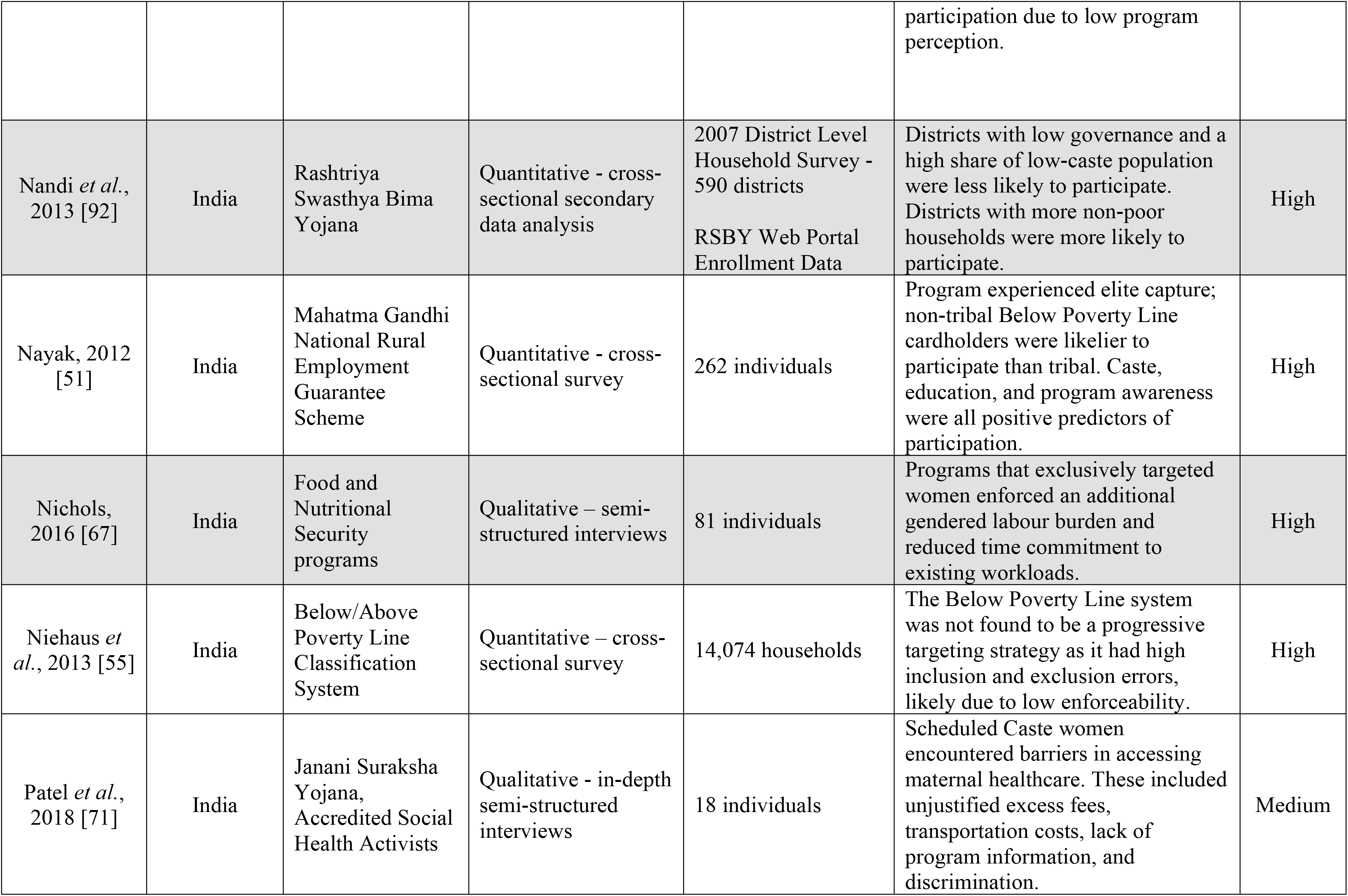
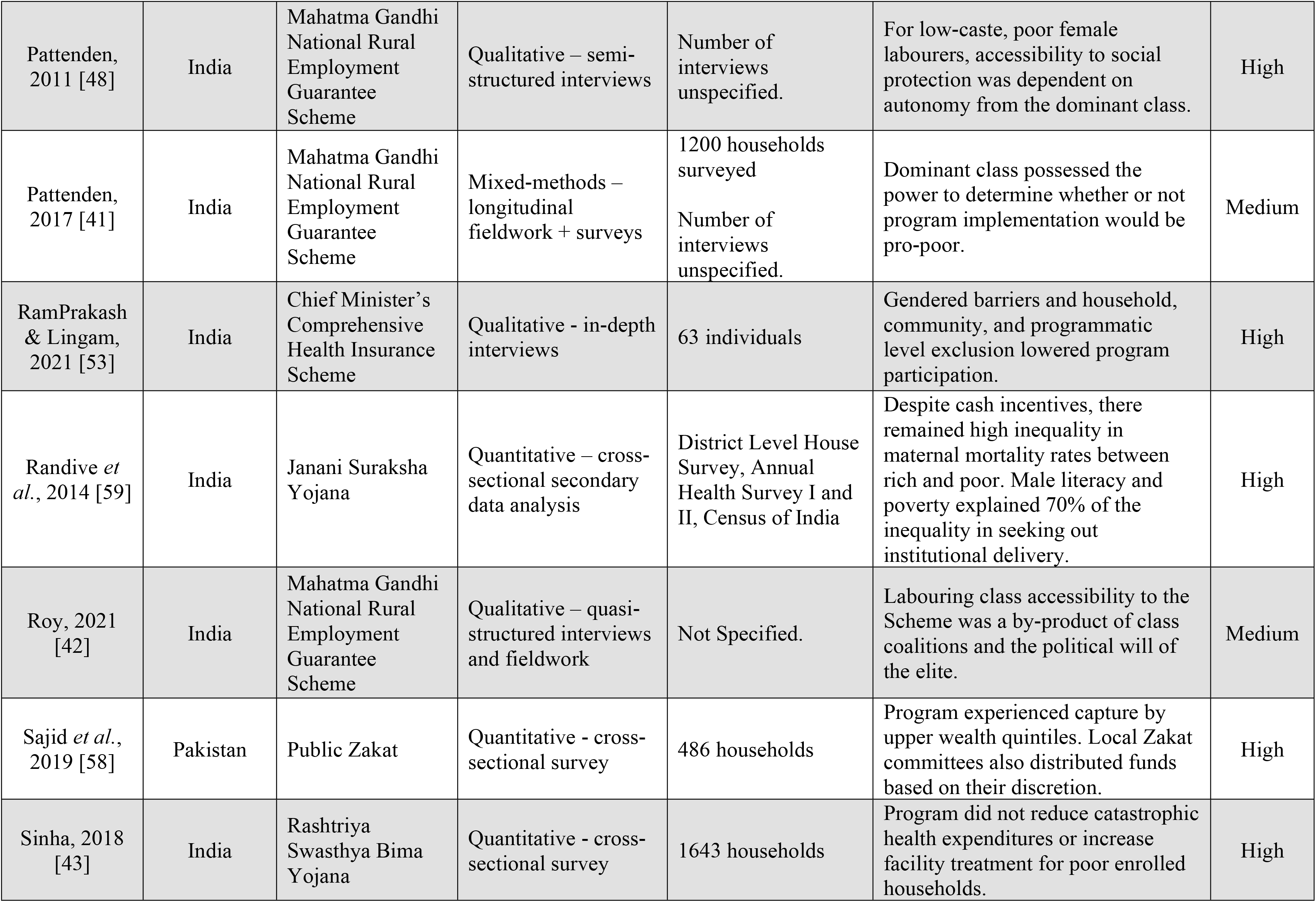
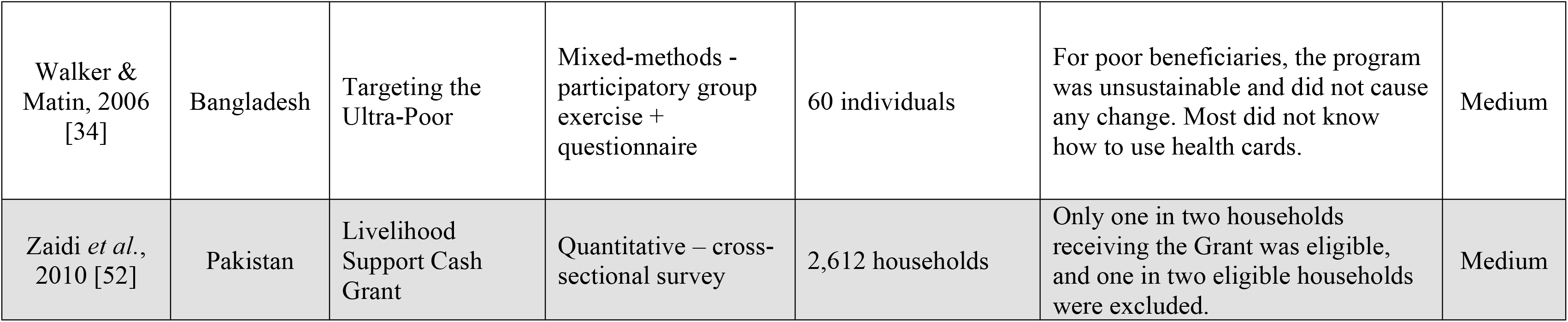
Summary of Reviewed Literature.

Data analysis was guided by Thomas & Harden’s [29] thematic synthesis approach. We first read the papers in detail and conducted a line-by-line coding. The codes were classified to develop descriptive categories. We then proceeded to conduct axial coding and inductive reasoning to develop broader analytical themes across the 42 papers. The iterative and data-driven approach allowed us to synthesize a higher-order interpretation of the literature as opposed to simply summarizing the findings. Importantly, this is not an exhaustive review but rather an analytical and thematic interpretation of the narrative surrounding South Asian social protection programs.

## Results

The articles in our review evaluate social protection initiatives across four countries - Bangladesh, India, Nepal, and Pakistan - collectively comprising over 95% of the South Asian population [30]. We identified 27 studies from India, seven from Pakistan, six from Bangladesh, and two from Nepal. Twenty-seven studies drew upon quantitative data, 11 used qualitative data, and four used both. The 42 papers researched a total of 23 programs. Table 4 summarizes these programs by country.

**Table 4.** Summary of Identified Social Protection Programs. *.

| Country | Program | Type of program | Eligible population | Program ongoing | Program summary | Literature |
| --- | --- | --- | --- | --- | --- | --- |
| Bangladesh | Challenging the Frontiers of Poverty-Targeting the Ultra-Poor | Microfinance | Households identified as ultra-poor who meet criteria | Yes | The program aims to help participants graduate from poverty by engaging them in sustainable entrepreneurial activities through education, training, and small loans. Launched in 2002, it targets poor, often landless, women earning less than \$0.60-\$0.70/day. | [33-35] |
|  | Food-for-Education | School enrollment & food subsidy | Poor children in public school | Yes** | Introduced in 1993, the program provided a free ration of grains to school children from poor families, to either consume or sell [93]. Households are targeted by community members who are also responsible for the distribution of entitlements (15kg for one child and 20kg for more than one child). | [44,91] |
|  | Shasthyo Shuroksha Karmasuchi | Healthcare subsidy | Below Poverty Line households | Yes | A pilot project aiming to reduce catastrophic out-of-pocket healthcare expenditures for poor households. The program covers inpatient healthcare services for 78 disease conditions at registered health facilities in three *upzilas* (Kalihati, Madhupar, and Ghatail). | [77] |
| India | Chief Minister's Comprehensive Health Insurance Scheme | Health Insurance | Below Poverty Line households | Yes | The program is a public-private partnership that covers medical and surgical treatment in empanelled hospitals in the state of Tamil Nadu. Eligible households must have an annual income of less than INR 72000, as proved by a ration card and income certificate. | [53] |
|  | Food & Nutritional Security Programs | Food security | N/A | N/A | A basket of programs aimed at addressing malnutrition by providing food rations, with a focus on mothers and children. It includes the Targeted Public Distribution System, Anganwadi Centres, Auxiliary Nurses and Midwives, and Accredited Social Health Activists. | [67] |
|  | Integrated Rural Development Programme | Microfinance | Below Poverty Line households | Yes** | First launched nationally in 1980, the program provides government subsidies and low-interest credit to poor households. It has now been merged with the Swarnajayanti Gram Swarozgar Yojana initiative, a microfinance program. | [69,89] |
| | Janani Suraksha Yojana | Cash incentive | Pregnant women in states with low institutional delivery | Yes | A cash-incentive to encourage women to give birth in a health facility. Rural women are given \$31 and urban women \$22. | [59,71] |
|  | Jawahar Rozgar Yojana | Employment | Rural men and women | Yes** | An employment generation program. Projects are determined by village *Gram Panchayat* members, with priority given to the development of rural infrastructure. | [69] |
|  | Mahatma Gandhi National Rural Employment Guarantee Scheme | Employment | Rural men and women | Yes | Introduced in 2005, the Scheme is a flagship initiative based on a constitutional commitment to provide 100 days of public work to rural households at statutory minimum wage. Rural men and women interested in participating apply through their local *Gram Panchayats*. There they are given job cards and can officially request work by filing written applications; if they are not given employment within two weeks, they are entitled to compensation via unemployment allowance. It is also mandatory that 33% of participants be women. | [39-42,45-48,51] |
|  | National Old Age Pension Scheme | Pension | Below Poverty Line aged 60+ | Yes | Old age beneficiaries receive 200 INR / month till the age of 79, after which they receive 500 INR / month [94]. | [56,62] |
|  | Rashtriya Swasthya Bima Yojana | Health insurance | Below Poverty Line households, and other disadvantaged groups | Yes | Introduced in 2008 as a cushion for poor households to avoid falling into poverty from catastrophic health expenditures. For an annual registration fee of 30 INR, the program covers up to 30,000 INR of inpatient treatment for five household members per year. | [43,92] |
|  | Rural Public Works | Employment | Rural men and women | Yes** | Employment generation program intended to develop rural infrastructure and provide work during agricultural slack periods. | [69,90] |
|  | Swarnajayanti Gram Swarozgar Yojana | Microfinance | Below Poverty Line households | Yes | A microcredit initiative that offers loans to support business development for women from Below Poverty Line households. Through either self-selection or the support of local leaders and organizations, participants are allocated into self-help groups where each member must contribute regularly to the outstanding loan. Through progressive lending, borrowers must pass a series of liability tests to become eligible for a greater amount of credit. | [61] |
|  | Targeted Public Distribution System | Food subsidy | Below Poverty Line and Antyodaya Anna Yojana households | Yes | In 1997, the Public Distribution became a targeted scheme that sells subsidized grains, sugar, oil, and kerosene through government-owned Fair Price Shops. The program was restricted to Below Poverty Line and Antyodaya Anna Yojana households, who were entitled to 35kg of food grains per month, with the latter group having an additional subsidy as they are categorized as the poorest of the poor. | [50,54,60,63,64] |
| **Nepal** | Dalit Child Grant | Cash transfer | Poor Dalit children under 5 years of age | Yes | Aims to address food insecurity and nutritional deficits by offering Dalit households 200 NPR per month for a maximum of two children under the age of 5 for five years. | [49] |
|  | Food-for-Work | Employment & food subsidy | Food insecure villages | Yes | Operating under the World Food Program, the initiative aims to improve food security while building rural infrastructure, focusing on agricultural development. | [37] |
|  | Nepal Food Corporation | Food subsidy | Food insecure districts | Yes | Introduced by the government in 1974, the Corporation provides subsidized grains to 30 remote and food insecure districts. Households are allocated 5 kg of rice per person per month. | [37] |
|  | Single Women Allowance | Cash transfer | Widows or single women aged 60+ | Yes | Acting as a form of social security allowance, the grant provides single women with a monthly transfer of 500 NPR. | [49] |
| **Pakistan** | Benazir Income Support Programme | Unconditional cash transfer | Ever-married women aged 18+ who meet eligibility criteria | Yes** | Beneficiaries are paid in quarterly installments of 5,000 PKR [95]. Eligibility criteria include being female, monthly income less than PKR 6,000, no family member in government service, and not being a beneficiary of any other program [96]. | [70] |
|  | Community Midwife Program | Healthcare services | Rural women | Yes | Adopted in 2006, the midwife program trains girls in skilled birth attendance to provide maternity care in rural villages. Midwives are expected to attract fee-paying clients by establishing a private practice. | [38] |
|  | Lady Health Workers | Healthcare services | Individuals in rural and urban slums | Yes | Community health worker program aimed at providing domiciliary primary healthcare services with a focus on maternal and child care. | [65,68,70] |
|  | Livelihood Support Cash Grant | Cash transfer | Eligible households in earthquake-affected areas | No | Introduced following the 2005 earthquake to provide affected households with 6 monthly installments of 3,000 PKR [97]. Beneficiary households must meet criteria such as severe housing damage, no male members aged 18-60, and/or low-ranking or no government officers. | [52] |
|  | Zakat Program | Unconditional cash transfer | Poor households | Yes | A tax-financed social transfer program based on the Islamic principle of charity. Funds are distributed by Local Zakat Committees, which are also responsible for identifying the poor. | [57,58] |

\* Summaries are provided according to the program design at the time of the most recent paper included in the review.
\*\* Program has been re-named, merged with another initiative, or modified.

Our analysis identified five themes underscoring the constructs and processes implicated in withholding social protection programs from reaching the poorest of the poor. These are summarized in Figure 2.

**Figure 2.**
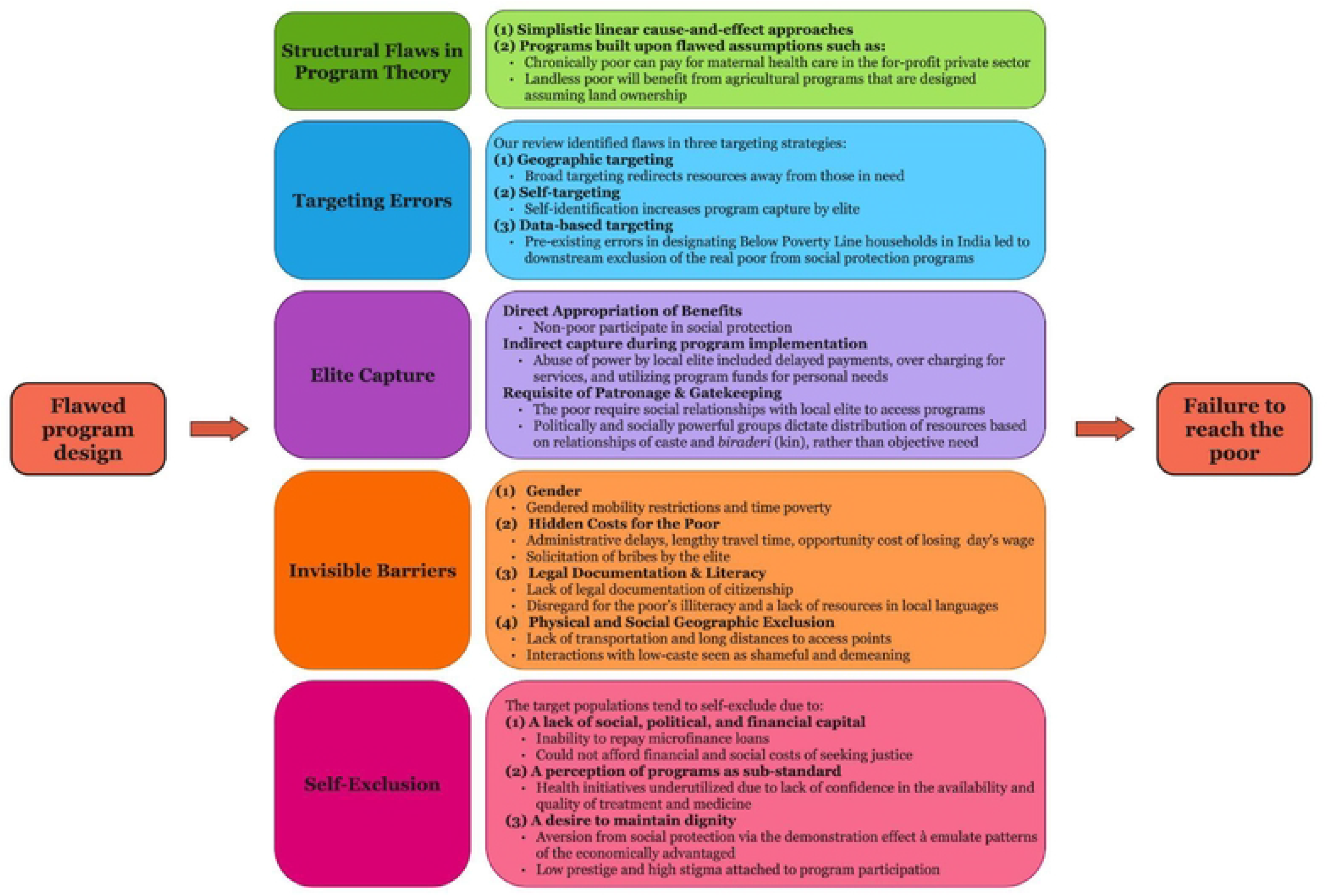
Summary of five themes inhibiting social protection programs from realizing their full potential in reaching the poor.

### Structural Flaws in Program Theory

Our review indicates that most programs had flawed program theory which outlines how an initiative will produce its intended outcomes (i.e., the program design) [31]. Program theories frequently rely on oversimplified linear models of cause and effect and presuppose that their one-dimensional initiatives will produce a chain of consequential impacts. They fail to consider the multi-dimensionality of poverty and how it is rooted in the broader social, economic, and political structures that adversely shape incorporation into society. This is best exemplified by Bangladesh’s Challenging the Frontiers of Poverty Reduction: Targeting the Ultra-Poor initiative, which aimed to redirect the destitute from low-skill occupations through education, income generation training, and microfinance loan services [32,33]. However, evaluations of the program showed that the landless households experienced negligible gains from participation [34]. Entrepreneurial activity increased in the short- and medium-term following program graduation, but the results were not sustainable in the long term [33,35]. Most beneficiaries returned to their baseline occupations, often as beggars and maids [33,35]. We argue this happened because the program’s design assumed that poverty is transient and that a one-time offer of micro-entrepreneurship support was sufficient to matriculate from penury [36]. It ignored the multiple forces operating to keep the poor poor, including their systematic exclusion from markets and the social relationships between and within networks that underpin entrepreneurial activities.

Another example of program theory deficiencies is the Nepal Food Corporation, which provides subsidised rice for the poor. The program required the beneficiaries collect the rice from central depots, which are located at significant distances from the remote villages where the target beneficiaries live [37]. Since the destitute lack the financial capital to travel to the central depots, the gap in service is filled by individuals with the means to purchase and transport large quantities of grains for the entire village. These middlemen sell the government-subsidized rice for a hefty profit - citing ‘transportation costs’ - forcing the poor to purchase their entitled rice quotas through high-interest credit. Consequently, the poor, low-caste Dalits only have 2% of their food needs met through the social protection program [37]. This crucial flaw in program design minimizes the program’s impact on food insecurity, instead nurturing profits for the rich and poverty for the poor.

Other program theories held built-in assumptions that inherently required that beneficiaries have a certain degree of wealth. The Community Midwife Program in Pakistan serves as an illustrative example. This program aimed to ensure skilled birth attendance for impoverished rural women. However, its placement within the private sector proved to be a fundamental flaw [38]. Cognizant of the dissonance between providing a service for the poor, while locating the midwives in the private sector, program managers suggested midwives charge a modest fee of USD 0.66 USD for childbirth. Yet, to maintain their practices, the midwives needed to charge fees 10 to 20 times higher than recommended. This private-sector-based approach ultimately failed to meet its objectives, imposing unrealistic financial burdens on both the impoverished women it aimed to serve and the midwives themselves. Over-estimation of the poor’s assets was also evident in Nepal’s Food-for-Work initiative, which has three-fold objectives: (1) improve agricultural output; (2) build road infrastructure to increase market accessibility; (3) provide poor households with employment opportunities by working on the farms or building infrastructure, for which they are paid in-kind with rice [37]. In the district of Humla, the program only benefitted the high-caste, rich landowners since they got cheap agricultural labour and road access from their lands to markets. Meanwhile, the landless, chronically poor - the key target group - got nothing more than an insufficient amount of rice, meeting only 15% of a household’s food requirements [37]. The initiative is essentially futile and trivial for the poor, for it failed to achieve its stated objectives of reducing food insecurity and poverty.

### Elite Capture of Social Protection Programs

The second theme emerging in our review was the concentration of power in the hands of the South Asian elite and how this impeded program reach to the poorest of the poor. In this context, the denomination of the elite is based on their status as high-caste, with its associated opportunities, privileges, and entitlements [37,39–42]. This includes land ownership, wealth, and membership in networks that sustain their dominant social and political identities.

### Direct appropriation of benefits by the elite

The first pattern of elite capture is a traditional form of corruption in which the non-poor participate in welfare programs meant for the poor. For instance, in their sample, Sinha [43] found that 47% of households enrolled in India’s Rashtriya Swasthya Bima Yojana health insurance scheme were non-poor. In Nepal and Bangladesh, the elite captured, or in simpler terms stole, subsidized grains meant for the food-insecure poor. Through secret transactions with program managers in Nepal’s Food Corporation program, large quantities of rice meant for the poor were diverted to local hotel owners to brew alcohol [37]. Bangladesh’s Food-for-Education program encountered a similar appropriation of grains. Despite providing medium-quality rice to disincentivize wealthy households, the rich still captured the resource - not for their consumption, but rather as a payment source to their poor servants [44]. The landed rich also captured India’s Mahatma Gandhi National Rural Employment Guarantee Scheme, a program designed to employ the rural poor. Those in the highest wealth quintile not only took over the employment opportunities meant for the landless, the illiterate, and the poor, but they also earned a higher daily wage [39,41,45–48]. Over 25% of poor households who sought work did not receive it despite being ‘guaranteed’ 100 days of work [45].

### Elite’s capture of resources through their power over program implementation

The elite also capture resources via the power they hold as social protection program implementers. Most program managers are the local elite since they meet the educational requirements and have the social capital needed to access such job opportunities. This role gives the traditional, informally powerful elite formal power. It simultaneously situates the poor at the mercy of the very people responsible for their subordination. In Nepal, personnel responsible for distributing cash transfers to the poor under the Dalit Child Grant delayed payments, instead redirecting funds into their personal accounts to collect interest [49]. Likewise, the elite captured the implementation of India’s Mahatma Gandhi National Rural Employment Guarantee Scheme. The program aimed to build village infrastructure and simultaneously provide employment opportunities for rural men and women [41,42]. The program planners designated the *Gram Panchayat,* a village-level governing council, as the implementing body responsible for all program tasks, including employment of labourers and allocation of work hours [41,42]. *Gram Panchayat* are traditionally made up of land affluent dominant classes. Many evaluations noted that the *Gram Panchayat* members captured the government-funded labour sources for their personal farms and other private projects [41,42]. For example, Pattenden [41] found that in the village of Panchnagaram, the appropriation of labour led to a third of the local program funding being allocated towards projects on the private lands of the *Gram Panchayat* members. These elites also disincentivized the poor from applying to the program by delaying and withholding wages, sharing misinformation, requiring hidden fees, hiring through nepotism, and, in some cases, preventing registration by withholding applications [39,41,42,46,48]. Ultimately, rather than reducing poverty, the program reproduced and even strengthened the dynamics of traditional inequity [41].

### The necessity of elite patronage & their gatekeeping role

The literature highlights how the elite often act as gatekeepers, controlling access to social protection for the poor. Here lies the catch-22 of social protection: it is supposed to offer an opportunity for the destitute to lower their dependency and potentially disengage from abusive relationships with the elite, but program design ensures it is precisely these relationships that the poor require to access the programs. Ideally, program accessibility should be based on state-citizen social contracts, but in South Asia, this ideal reigns inferior to the informal contracts with local elites [37,39]. The politically and socially powerful groups dictate the distribution of resources based on caste and *biraderi* (kin) relationships [38,39,50–52]. Due to their high decision-making entitlement and low accountability, they act as the poor’s primary route to access social protection [39,53]. In India, these bonds between the powerful and poor are so integral to societal structure that the practice of their formation has been given the special name of *bhav-vyavhar*, or the emotional and behavioural relationship building with the elite [39]. *Bhav-vyavhar* occurs through social transactions such as the poor offering free physical labour, emotional support, and even financial transfers, including a portion of their pension.

In India and Pakistan, some program designs allocated the task of identifying poor households to municipal-level officials and the village elite (often the same group). These officials registered their non-poor family and friends instead of the destitute [54,55]. In India, the poor perceived India’s National Old Age Pension Scheme as unequal and inaccessible since they lacked the social linkages necessary for participation in the program [56]. This is best reflected in the finding that 74% of deserving recipients were not receiving the Pension [56]. Gatekeeping was also operative in Pakistan’s Zakat initiative, in which Local Zakat Committees, comprised of the community’s well-off, are responsible for identifying the poor and eligible households [57]. An evaluation by Sajid *et al*. [58] showed that in 2019, 40% of the beneficiaries were non-poor.

A powerful tool is the elite’s control of knowledge about social programs’ existence and benefits. Most program designs place the responsibility of disseminating information in the hands of the elite, like the Local Zakat Committees in Pakistan. An evaluation [58] found that the Committees were withholding this knowledge as 80% of eligible beneficiaries in their sample were unaware of the benefits they were entitled to. Das *et al.* [46] also reported censorship of information in their evaluation of India’s Mahatma Gandhi National Rural Employment Guarantee Scheme. The elite forming the *Gram Panchayats* withheld information to ensure local villagers remained unaware of the basic guidelines and operations of the program. Consequently, the poor beneficiaries were underpaid and underemployed.

### Targeting Errors

The third theme in our review was the insensitivity of program design when targeting beneficiaries. The literature identified three targeting strategies traditionally used to reach recipients: geographic targeting, self-targeting, and data-based targeting. Geographic targeting purposively selects regions based on poverty or health indicators. Self-targeting (or self-selection) places the onus of identifying eligibility on the poor themselves. Data-based targeting relies on a pre-existing national dataset, such as India’s Below Poverty Line classification. Our review showed that all three mechanisms failed to target their ideal beneficiaries sufficiently and were instead more likely to reach undeserving households.

The Janani Suraksha Yojana program in India illustrates the insensitivity of geographic targeting. This initiative specifically focused on states with low rates of institutional deliveries, introducing a cash incentive scheme to encourage women to deliver in healthcare facilities. [59]. An evaluation in nine participating states revealed that all socioeconomic groups experienced a similar average increase in institutional birth. However, the reduction in maternal mortality was four times greater for the richest quintile compared to the poorest [59]. Thus, despite a principal program objective being to lessen financial barriers for *poor* women, the broad geographic targeting mechanism disproportionately benefited the rich living in these localities.

India’s Mahatma Gandhi National Rural Employment Guarantee Scheme serves as a prime example of self-targeting. The program provides employment opportunities for unskilled labourers [47]. An evaluation [47] showed self-targeting permitted uptake by all socioeconomic groups, irrespective of need. In the state of Tamil Nadu, only 40% of the participants were truly poor. Likewise, Pakistan’s Livelihood Support Cash Grant implemented a similarly ineffective form of self-targeting. Applications for the Grant were open to all, but applicants were subject to a screening process based on the government’s predetermined eligibility criteria [52]. Although the program enrolled nearly 700,000 families as beneficiaries, a community-based survey showed that half of the eligible families were excluded, while half of the ineligible rich were provided benefits [52].

A widely recognized static targeting strategy in India is based on classifying households as Below or Above Poverty Line through a state-level census. This classification is not infallible as an evaluation found that 70% of ineligible households in the state of Karnataka possessed Below Poverty Line cards, while 13% of eligible households did not [55]. The downstream consequences of this targeting flaw become evident in various social programs’ failure to reach the poor [55,60]. One example is the failure of Swarnajayanti Gram Swarozgar Yojana microfinance program for the destitute. An evaluation [61] found that none of the beneficiaries in their sample were extremely poor, while over half of the program participants were from high-income households falsely possessing Below Poverty Line cards. Another example is the National Old Age Pension Scheme, in which non-poor Below Poverty Line cardholders had a higher likelihood of receiving the social allowance than poor cardholders [56]. In fact, Mishra & Kar [62] found that 50% of the Pension recipients were from the top two income quartiles. A third example can be found in India’s Targeted Public Distribution System, a food ration initiative. Five evaluations [50,54,60,63,64] found that a large fraction of the wrongly classified and wealthy Below Poverty Line cardholders were receiving food rations, while a substantial proportion of the poorest households were not covered by the program. These are worrying findings given that India’s poor lead a dangerous “hand-to-mouth existence” [50].

### Invisible and Unacknowledged Barriers

Program designs failed to take into account a host of barriers handicapping the poor’s accessibility to social protection. These include gendered barriers, hidden costs, the poor’s lack of legal documentation, and the geographies of exclusion. We discuss each below.

### Gendered barriers

Despite decades of advocacy, programs remain gender blind. In Pakistan, programs like the Lady Health Worker and Community Midwife are specifically designed to deliver domiciliary health care services to home-bound women in direct acknowledgment of the cultural practice of *pardah* (seclusion), and its’ associated value of male honor, limiting women’s unaccompanied movement [38,65,66]. The same gender values also require that the health workers be female.

Consequently, only women are recruited as Lady Health Workers and Midwives to provide home-based services. In doing so, the program design paradoxically expects the female health workers to transgress the very gender norms that created the need for their services in the first place. A reading of the papers suggests the program designers were aware of women’s gendered mobility restrictions but chose to ignore the implications of this reality for the female community health workers. This insensitivity and lack of consideration for the young Lady Health Workers and Midwives’ safety and respectability discouraged the workers from assisting households beyond a short walking distance or within their social geography [38,65,66]. The most vulnerable women remained underserved.

Similarly, in Nepal, a food security intervention reproduced gendered inequalities by erring in assuming female labour was a freely available commodity [67]. The program activities added to women’s existing time-consuming socially ascribed domestic duties. This led to female time poverty and subsequent selectivity in program participation [67]. The most effective development strategies were those that relieved gendered labour burdens through services such as the provision of school lunches [67].

### Hidden costs for the poor

Another crucial structural flaw in program design is the hidden costs required to access the programs. One such cost is the time needed to travel and fulfill the often-onerous bureaucratic requirements, which often results in the loss of a full day’s wage [53,68]. An evaluation of India’s Targeted Public Distribution System in the sub-district of Sasan found that 64% of households without ration cards did not even apply out of concern that administrative delays would interfere with work schedules and subsequent earnings [64]. Given the poor’s fragility of income, these opportunity costs can considerably compound the depths of their poverty - resulting in a net financial loss rather than a benefit.

The assumption that time is trivial for the destitute was also exemplified in the design flaws within Pakistan’s Livelihood Support Cash Grants program [52]. To find out if they had been accepted as beneficiaries, poverty-stricken victims of a natural disaster were expected to make regular trips to district offices. If denied, families could only contest their eligibility status within the first seven days following the publication of recipient lists, which were released on an unpredictable schedule. Without regular travel to district offices, poor households could miss the opportunity to appeal - likely a contributing factor to the finding that half of the eligible beneficiaries were not reached by the program [52].

Another hidden cost is the solicitation of bribes. An overwhelmingly common phenomenon within South Asian social protection, corruption is often legitimized under the guise of registration fees and gratuities [39,46,50,69–71]. Taking the example of Pakistan’s Zakat cash transfers, the monetary value is reduced as beneficiaries highlight multiple hidden transactional fees from registration officers, peons, and even postmen [57]. In other cases, applicants were completely unaware that they were paying bribes - believing it to be a requisite for program participation [49].

### Lack of legal documentation and literacy

One common but truly invisible barrier was the poor’s exclusion from documentation of their legal citizenship. Program designs nearly always require such documentation, often an ID card or birth certificate [50,53,56,57,71]. The poor lack these documents for a range of reasons including but not limited to the lack of a permanent address [72], lack of knowledge of the importance of documentation [73], the cost of obtaining documentation [73–75], internal displacement [76], and generational systemic exclusion due to gender [73,74] or caste [75]. These groups contribute to the ‘Invisible Billion’ worldwide who cannot prove their legal identity [74]. In India, the poor’s lack of appropriate documentation meant they could not apply for Below Poverty Line ration cards, which were needed to prove poverty status when registering for the Chief Minister’s Comprehensive Health Insurance Scheme [53]. Some applicants were also excluded from the country’s Rashtriya Swasthya Bima Yojana health insurance as their presented records did not match those of the company handling enrollment [43]. For Pakistan’s Zakat program, these requirements created a black market for illegal birth certificates [57].

Social protection programs also faltered in accommodating the close relationship between poverty and low literacy in rural South Asia [46,52]. Program resources were provided in written format, often in English. The poor’s subsequent inability to absorb the information reified relationships of dependency on the local elite and government staff [46,52].

### Geography of exclusion

Geographic exclusion encompasses social and physical dimensions, an aspect that social protection programs often overlook in program designs. Despite its seemingly obvious role in accessibility, eight initiatives failed to consider how the lack of transportation and long distances to access points might limit uptake by the poor [34,38,43,53,68,71,77]. The program designs of India’s Targeted Public Distribution System and the Nepal Food Corporation required the destitute to travel to far, often centrally-located, ration shops at their own expense, incurring high transaction costs [37,54,63,64]. In Nepal, Gautam & Andersen [37] found that households with limited mobility resources would spend days walking through harsh terrain to procure their grain rations. Their travel burden was worsened by the uncertainty of whether or not depots would be open or even be capable of meeting their quotas. The program design ultimately penalized the most marginalized, the poor living in remote villages.

Social geography’s less obvious (but critical) role was overlooked in the design of Pakistan’s Lady Health Worker program [65]. Women living in *bhattas* (brick kilns), encompassing the poorest of the poor populations, were unreached by the program because the *bhattas* were considered no-go zones and not a part of human settlement. They were, consequently, beyond the borders of the workers’ catchment areas [70]. There was also the perception that it was “demeaning” to enter *bhattas* [70].

### Gateways to Self-Exclusion by the Poor

Our final theme is the poor choosing to self-exclude from participating in social protection programs. We identified three possible gateways for this process: a desire to maintain dignity, a lack of capital, and a perception of programs as substandard.

The first form of self-exclusion is a desire to maintain a sense of dignity. This is best described by Axel Honneth [78], who says, “We owe our integrity… to the receipt of approval or recognition from other persons.” Nair [54] introduces the idea of aversion to social protection via the demonstration effect in which poor households emulate the patterns of the economically advantaged. This occurred following India’s Public Distribution System’s transition from a universal to a targeted program. As Above Poverty Line households were no longer eligible, low prestige was assigned to those who continued participating in the ration system. Participation stigma was also evident in India’s Mahatma Gandhi National Rural Employment Guarantee Scheme, in which both tribal and non-tribal families considered the work as below their dignity [51]. Beneficiaries of Pakistan’s Zakat program complained about the demeaning application process; they felt the government treated them like “beggars” to receive a mere 200 PKR a month [57]. The gravity of aid-affiliated shame is further demonstrated by a village in northern Punjab, Pakistan, in which some poor low-caste women actually preferred to remain invisible and untargeted by social protection to avoid attracting unnecessary attention to their already disgraced identities [70].

The second form of self-exclusion results from the poor lacking the financial, social, and political capital often required to access the resources ostensibly for them. A poverty-stricken household’s awareness of its externally - and sometimes internally - imposed limitations can restrict it from even attempting to access social protection programs. This is evident in the poor’s hesitancy to participate in India’s Swarnajayanti Gram Swarozgar Yojana microfinance initiative due to their fear of being unable to repay their loans [61]. Lack of capital also translated into the sometimes insurmountable challenges of seeking *sunvaiyi* (justice) from government officials [39]. Although the poor had warranted complaints of elite capture, receipt of justice required financial input and high social status to ensure that their grievances would not be disregarded.

The last gateway to self-exclusion is a consequence of the perception that programs are of low quality. For example, despite recognizing the benefits of skilled maternal care, Pakistan’s low-caste women continued to seek unskilled birth attendants to avoid experiencing inferior treatment - mitigating the value of government-funded healthcare [70]. In India’s Chief Minister’s Comprehensive Health Insurance Scheme, access to funds took up to 25 days, which delayed treatment even during emergencies [53]. Poor women instead subjected themselves to out-of-pocket payments to receive immediate and poor-quality care. Villagers’ perceptions of programs were also guided by bad experiences of seeking care shared within the community, thus deterring some ideal beneficiaries, and fabricating a form of collective exclusion [68].

## Discussion

While our aim is not to belittle the positive outcomes of social protection, the principal conclusion of our review of the literature is that the root cause of why programs fail to reach the poor lie in program designs that are disconnected from on-the-ground realities. This is not a new discovery. Many authors have commented on this disconnect and have highlighted it as the underlying reason for why programs are not truly transformative [79]. This begs the question, why does this apathy towards the ground realities of the chronically poor persist?

We propose there are three reasons for this. First: there is a lack of understanding of who precisely the poor are in South Asia. Despite a vast body of literature describing the region’s caste system, it has not been incorporated in national-level definitions of the poor. All criteria for capturing poverty continue to be determined by western-led definitions [80]. Second: despite the rhetoric of co-production and end-user engagement [81], there remains a gap in our knowledge on how to precisely engage with communities at large and especially with poor communities [82]. Third: the structures and timelines of funding calls do not lend themselves to any meaningful engagement with poor end-users. Despite requiring engagement with ‘local country stakeholders’, donors give 4-6 weeks to develop the program proposals. For example, a recent multi-million-dollar funding call from Global Affairs Canada, ‘Resilient Health Systems for All’ [83], gave 6 weeks for project design, development of relationships with international stakeholders and write-up. Clearly, there was no time to engage with the primary stakeholders of these projects, the poor. We suggest that the disconnect between program design and poor’s ground reality is intentional and reflects a broader power dynamic. It represents the global elite’s effort to preserve their dominance, with wealthy donor nations and local elites dictating the lives of poorer populations based on geopolitical interests [84] and national priorities [85].

The review had some limitations. The first essentially reflects the limitations of the literature. Twenty-seven out of the 42 studies included in our review were quantitative surveys, a rigid data collection method contingent on the quality of questions and their responses [86]. Quantitative results fail to capture behavioural and emotional attributes from respondents, both valuable measures when evaluating the socially and politically driven accessibility barriers encountered by the poor [86]. Second, when identifying papers from outside the databases, we potentially subjected ourselves to the bias of including studies supporting our themes. Third, our review did not capture data from all South Asian countries as there is a paucity of literature describing and evaluating social protection programs in Afghanistan and Sri Lanka.

Based on this review of the literature, we recommend the traditional approaches to designing and implementing social protection programs need to be overhauled and decolonized. This needs to start from the structures of donor funding seeding program ideas, and follow-through to national governments that carry the programs forward. There is a need to critically appraise the international consultants and local decision-makers designing these programs in terms of their contextual understanding and accountability. We also recommend more research to understand who the truly poor are and how to best engage them in program design. This requires greater use of qualitative and other innovative research methods. In-depth studies should also be conducted to sociologically evaluate the influence of the caste system on social protection programs, especially in the Muslim-dominant countries of Pakistan and Bangladesh [20,87,88]. The research should contextualize how the power dynamics created by this system are at play in the social, economic, and political spheres and, consequently, must be accounted for in the design of social protection programs. Lastly, research must be conducted to identify how South Asian governments can be held accountable to their poor citizens, and effectively absorb the existing knowledge.

## Data Availability

This submission is a literature review. We did not use any empirical data and therefore there is no data for sharing

## Acknowledgements

The authors thank all researchers involved in the original projects.

## Notes

### Competing Interest Statement

The authors have declared no competing interest.

### Funding Statement

The authors received no specific funding for this work.

### Author Declarations

Alberta Research Information Services System (ARISE) - University of Alberta No: PRO 00117838

## References

1. Bloch C. Social spending in South Asia— an overview of government expenditure on health, education and social assistance. Brasília and Katmandu: International Policy Centre for Inclusive Growth; 2020. 57 p. Report No.:44.

2. Mitra A, Paul M. Poverty targeting and economic growth in India. J Income Wealth. 2018;40(2):177–89.

3. Kharas H, Dooley M. Extreme poverty in the time of COVID-19. Washington (DC): Center for Sustainable Development at Brookings; 2021 Jun. 11 p.

4. Kabeer N. Scoping study on social protection: evidence on impacts and future research directions. [place unknown]: DFID Research and Evidence; 2009 Dec. 67 p.

5. Kabeer N. The politics and practicalities of universalism: towards a citizen-centered perspective on social protection. Eur J Dev Res. 2014 Jul 1;26:338–54.

6. OECD Development Centre. (2019). Lessons from the EU-SPS programme: implementing social protection strategies. Paris (FR): OECD; 2019. 41 p.

7. Dreze J, Sen A. Public action for social security: foundations and strategy. In: Ahmed E, Dreze J, Hills J, Sen A, editors. Social security in developing countries. New York (NY): Oxford University Press; 1991. p. 2–40.

8. Guhan S. Social security options for developing countries. In: Figueiredo JB, Shaheed Z, editors. Reducing poverty through labour market policies. Geneva (CH): International Labour Office; 1994. p. 35–53.

9. Devereux S, McGregor JA. Transforming social protection: human wellbeing and social justice. Eur J Dev Res. 2014 Jul 1;26(3):296–310.

10. O’Brien C, Scott Z, Smith G, Barca V, Kardan A, Holmes R, Watson C, Congrave J. Shock-responsive social protection systems research: synthesis report. Oxford (UK): Oxford Policy Management; 2018 Jan. 104 p.

11. Carter B, Roelen K, Enfield S, Avis W. Social protection topic guide. Brighton (UK): Institute of Development Studies; 2019 Oct. 68 p.

12. United Nations. Secretary-General’s Policy Brief: Investing in jobs and social protection for poverty eradication and a sustainable recovery. [place unknown]: United Nations; 2021 Sep 28. 28 p.

13. Gentilini U. Cash transfers in pandemic times: evidence, practices, and implications from the largest scale up in history. Washington (DC): World Bank; 2022 Jul. 100 p.

14. De Schutter O. Non-take-up of rights in the context of social protection [Internet]. [place unknown]: United Nations Human Rights Council; 2022 Apr 19 [cited 2023 Sep 26]. Available from: https://documents-dds-ny.un.org/doc/UNDOC/GEN/G22/322/17/pdf/G2232217.pdf?OpenElement

15. Kidd S, Athias D. Hit and miss: an assessment of targeting effectiveness in social protection with additional analysis. London (UK): Development Pathways; 2020 Jun. 91 p.

16. Grosh M, Leite P, Wai-Poi M, Tesliuc E. Revisiting targeting in social assistance: a new look at old dilemmas. Washington (DC): World Bank; 2022.

17. Mahmood M. Global debt crisis and South Asia. The Financial Express [Internet]. 2022 Aug 13 [cited 2023 Sep 26]. Available from: https://thefinancialexpress.com.bd/views/views/global-debt-crisis-and-south-asia-1660399998

18. Devarajan S, Nabi I. Economic growth in South Asia: promising, unequalising, sustainable? Econ Political Wkly. 2006 Aug 19;41(33):3573–80.

19. World Bank Group. Climate change action plan 2021-2025: South Asia roadmap. Washington (DC): World Bank Group; 2021 [cited 2023 April 17]. 146 p. Available from: https://openknowledge.worldbank.org/bitstream/handle/10986/36321/164599.pdf

20. Basu, K. Why is Bangladesh booming? Brookings [Internet]. 2018 May 1 [cited 2023 Oct 31]. Available from: https://www.brookings.edu/articles/why-is-bangladesh-booming/#:~:text=Since%20then%2C%20Bangladesh%27s%20annual%20GDP,reversed%20barring%20gross%20policy%20mismanagement).

21. Thresia CU, Srinivas PN, Mohindra KS, Jagadeesan CK. The health of Indigenous populations in South Asia: a critical review in a critical time. International Journal of Health Services. 2022 Jan;52(1):61–72.

22. Bhutta ZA, Mitra A, Salman A, Akbari F, Dalil S, Jehan F, Chowdhury M, Jayasinghe S, Menon P, Nundy S, Qadri F. Conflict, extremism, resilience and peace in South Asia; can covid-19 provide a bridge for peace and rapprochement? BMJ. 2021 Nov 15;375.

23. World Economic Forum. Global gender gap report. Geneva (CH): World Economic Forum; 2023 Jun. 382 p.

24. Mumtaz Z, Jhangri GS, Bhatti A, Ellison GTH. Caste in Muslim Pakistan: a structural determinant of inequities in the uptake of maternal health services. Sex Reprod Health Matters. 2022;29(2):230–52.

25. Atlas of Social Protection Indicators of Resilience and Equity [Data set]. World Bank Group: Washington (DC); 2021 [cited 2023 April 17]. Available from: https://databank.worldbank.org/reports.aspx?source=1229#

26. Page MJ, McKenzie JE, Bossuyt PM, Boutron I, Hoffmann TC, Mulrow CD, et al. The PRISMA 2020 statement: an updated guideline for reporting systematic reviews. BMJ. 2021;372(n71):1-9.

27. What is GRADE? [Internet]. [place unknown]: GRADE; 2023 [cited 2023 Sep 26]. Available from: https://www.gradeworkinggroup.org/

28. Lewin S, Booth A, Glenton C, Munthe-Kaas H, Rashidian A, Wainwright M, et al. Applying GRADE-CERQual to qualitative evidence synthesis findings: introduction to the series. Implement Sci. 2018 Jan;13(1):1–10.

29. Thomas J, Harden A. Methods for the thematic synthesis of qualitative research in systematic reviews. BMC Med Res Methodol. 2008 Dec;8:1–10.

30. Population, total - India, Pakistan, Bangladesh, Nepal, South Asia [Data set]. World Bank: Washington (DC); 2021 [cited 2023 April 17]. Available from: https://data.worldbank.org/indicator/SP.POP.TOTL?locations=IN-PK-BD-NP-8S

31. Davidoff F, Dixon-Woods M, Leviton L, Michie S. Demystifying theory and its use in improvement. BMJ Qual Saf. 2015 Mar 1;24(3):228–38.

32. Hulme D, Moore K, Seraj KFB. Reaching the people whom microfinance cannot reach: Learning from BRAC’s “Targeting the Ultra Poor” programme. In: Armendariz B, Labie M, editors. The handbook of microfinance. Singapore: World Scientific Publishing Co. Pte. Ltd.; 2011. p. 563–86.

33. Misha FA, Raza WA, Ara J, Van de Poel E. How far does a big push really push? Long-term effects of an asset transfer program on employment trajectories. Econ Dev Cult Change. 2019 Oct 1;68(1):41–62.

34. Walker S, Matin I. Changes in the lives of the ultra poor: an exploratory study. Dev Pract. 2006 Feb 1;16(1):80–4.

35. Asadullah MN, Ara J. Evaluating the long-run impact of an innovative anti-poverty programme: evidence using household panel data. Appl Econ. 2016 Jan 8;48(2):107–20.

36. Sabates-Wheeler R. Graduation. In: Schüring E, Loewe M, editors. Handbook on social protection systems. Cheltenham (UK): Edward Elgar Publishing; 2021. p. 262–75.

37. Gautam Y, Andersen P. Aid or abyss? Food assistance programs (FAPs), food security and livelihoods in Humla, Nepal. Food Secur. 2017 Apr;9(2):227–38.

38. Mumtaz Z, Levay A, Bhatti A, Salway S. Good on paper: the gap between programme theory and real-world context in Pakistan’s Community Midwife programme. BJOG: Int J Obstet Gynaecol. 2015 Jan;122(2):249–58.

39. Akerkar S, Joshi PC, Fordham M. Cultures of entitlement and social protection: Evidence from flood prone Bahraich, Uttar Pradesh, India. World Dev. 2016 Oct 1;86:46–58.

40. Jha R, Bhattacharyya S, Gaiha R, Shankar S. “Capture” of anti-poverty programs: an analysis of the National Rural Employment Guarantee Program in India. J Asian Econ. 2009 Sep 1;20(4):456–64.

41. Pattenden J. Class and social policy: the National Rural Employment Guarantee Scheme in Karnataka, India. J Agrar Change. 2017 Jan;17(1):43–66.

42. Roy I. Class coalitions and social protection: the labouring classes and the National Rural Employment Guarantee Act in Eastern India. J Dev Stud. 2021 Jun 3;57(6):863–81.

43. Sinha RK. Impact of publicly financed health insurance scheme (Rashtriya Swasthya Bima Yojana) from equity and efficiency perspectives. Vikalpa. 2018 Dec;43(4):191–206.

44. Galasso E, Ravallion M. Decentralized targeting of an antipoverty program. J Public Econ. 2005 Apr 1;89(4):705–27.

45. Das U. Rationing and accuracy of targeting in India: the case of the Rural Employment Guarantee Act. Oxf Dev Stud. 2015 Jul 3;43(3):361–78.

46. Das U, Singh A, Mahanto N. Awareness about Mahatma Gandhi National Rural Employment Guarantee Act: some evidence from the northern parts of West Bengal, India. Econ Bull. 2012;32(1):528–37.

47. Jha R, Gaiha R, Shankar S, Pandey MK. Targeting accuracy of the NREG: evidence from Madhya Pradesh and Tamil Nadu. Eur J Dev Res. 2013 Dec 1;25(5):758–77.

48. Pattenden J. Social protection and class relations: evidence from scheduled caste women’s associations in rural South India. Dev Change. 2011 Mar;42(2):469–98.

49. Drucza K. Cash transfers in Nepal: Do they contribute to social inclusion? Oxf Dev Stud. 2016 Jan 2;44(1):49–69.

50. Khan GA. A critical analysis of vulnerability in informal sector employment in India: protective mechanisms and adequacy of protection. Int Soc Sci J. 2021 Sep;71(241-242):197–215.

51. Nayak S. What motivates to participate in an employment guarantee programme in India? A logit model analysis. Econ Bull. 2012;32(3):2113–27.

52. Zaidi S, Kamal A, Baig-Ansari N. Targeting vulnerability after the 2005 earthquake: Pakistan’s Livelihood Support Cash Grants programme. Disasters. 2010 Apr;34(2):380–401.

53. RamPrakash R, Lingam L. Why is women’s utilization of a publicly funded health insurance low? A qualitative study in Tamil Nadu, India. BMC Public Health. 2021 Feb 12;21:1–21.

54. Nair R. Public distribution system in Kerala reassessed. Indian J Soc Work. 2011 Jan;72(1):23–54.

55. Niehaus P, Atanassova A, Bertrand M, Mullainathan S. Targeting with agents. Am Econ J: Econ Policy. 2013 Feb 1;5(1):206–38.

56. Asri V. Targeting of social transfers: are India’s poor older people left behind? World Dev. 2019 Mar 1;115:46–63.

57. Kabeer N, Mumtaz K, Sayeed A. Beyond risk management: vulnerability, social protection and citizenship in Pakistan. J Int Dev. 2010 Jan 1;22(1):1–19.

58. Sajid GM, Arif GM, Yasin HM. Targeting efficiency of cash transfers programmes in Pakistan. Pakistan Econ Soc Rev. 2019 Jul 1;57(1):1–22.

59. Randive B, San Sebastian M, De Costa A, Lindholm L. Inequalities in institutional delivery uptake and maternal mortality reduction in the context of cash incentive program, Janani Suraksha Yojana: results from nine states in India. Soc Sci Med. 2014 Dec 1;123:1–6.

60. Mazumdar S, Sharma AN. Poverty and social protection in urban India: targeting efficiency and poverty impacts of the Targeted Public Distribution System. Indian J Labour Econ. 2013 Oct 1;56(4):547–77.

61. Mukherjee AK, Kundu A. Swarnajayanti Gram Swarojgar Yojona as a safety net: evidence from Murshidabad district of West Bengal. Bangladesh Dev Stud. 2012 Mar 1;35(1):79–103.

62. Mishra AK, Kar A. Are targeted unconditional cash transfers effective? Evidence from a poor region in India. Soc Indic Res. 2017 Jan;130(2):819–43.

63. Jha R, Gaiha R, Pandey MK, Kaicker N. Food subsidy, income transfer and the poor: a comparative analysis of the public distribution system in India’s states. J Policy Model. 2013 Nov 1;35(6):887–908.

64. Kannan KP, Pillai NV. Food security at the local level: a study in contrast between Kerala and Orissa in India. In: Cook S, Kabeer N, editors. Social protection as development policy: Asian perspectives. New Dehli (IN): Routledge; 2010. p. 245-277.

65. Mumtaz Z, Salway S, Nykiforuk C, Bhatti A, Ataullahjan A, Ayyalasomayajula B. The role of social geography on Lady Health Workers’ mobility and effectiveness in Pakistan. Soc Sci Med. 2013 Aug 1;91:48–57.

66. Mumtaz Z, Salway S. ‘I never go anywhere’: extricating the links between women’s mobility and uptake of reproductive health services in Pakistan. Soc Sci Med. 2005 Apr 1;60(8):1751–65.

67. Nichols CE. Time Ni Hota Hai: time poverty and food security in the Kumaon hills, India. Gend Place Cult. 2016 Oct 2;23(10):1404–19.

68. Bechange S, Schmidt E, Ruddock A, Khan IK, Gillani M, Roca A, et al. Understanding the role of lady health workers in improving access to eye health services in rural Pakistan–findings from a qualitative study. Arch Public Health. 2021 Dec;79(1):1–12.

69. Gaiha R, Imai K, Kaushik PD. On the targeting and cost-effectiveness of anti-poverty programmes in rural India. Dev Change. 2001 Mar;32(2):309–42.

70. Mumtaz Z, Salway S, Bhatti A, Shanner L, Zaman S, Laing L, et al. Improving maternal health in Pakistan: toward a deeper understanding of the social determinants of poor women’s access to maternal health services. Am J Public Health. 2014 Feb;104(S1):S17–24.

71. Patel P, Das M, Das U. The perceptions, health-seeking behaviours and access of scheduled caste women to maternal health services in Bihar, India. Reprod Health Matters. 2018 Nov 8;26(54):114–25.

72. Chandran R, Sharma S. Indian children without Aadhaar digital ID shut out of school [Internet]. [place unknown]: Context News; 2022 Jul 28 [cited 2023 Oct 31]. Available from: https://www.context.news/digital-divides/indian-children-without-aadhaar-digital-id-shut-out-of-school

73. Livani T, Haddock S. Women’s access to identification cards can accelerate development in Afghanistan [Internet]. [place unknown]: World Bank Blogs; 2020 Oct 5 [cited 2023 Oct 31]. Available from: https://blogs.worldbank.org/endpovertyinsouthasia/womens-access-identification-cards-can-accelerate-development-afghanistan

74. Desai VT, Diofasi A, Lu J. The global identification challenge: Who are the 1 billion people without proof of identity? [Internet]. [place unknown]: World Bank Blogs; 2018 Apr 25 [cited 2023 Oct 31]. Available from: https://blogs.worldbank.org/voices/global-identification-challenge-who-are-1-billion-people-without-proof-identity

75. Sur P. Under India’s caste system, Dalits are considered untouchable. The coronavirus is intensifying that slur [Internet]. [place unknown]: CNN; 2020 Apr 16 [cited 2023 Oct 31]. Available from: https://www.cnn.com/2020/04/15/asia/india-coronavirus-lower-castes-hnk-intl/index.html

76. Nayar R, Gottret P, Mitra P, Betcherman G, Lee YM, Santos I, Dahal M, Shrestha M. More and Better Jobs in South Asia. Washington (DC): World Bank; 2012.

77. Hasan MZ, Ahmed MW, Mehdi GG, Khan JA, Islam Z, Chowdhury ME, et al. Factors affecting the healthcare utilization from Shasthyo Suroksha Karmasuchi scheme among the below-poverty-line population in one subdistrict in Bangladesh: a cross sectional study. BMC Health Serv Res. 2022 Jul 8;22(1):885.

78. Honneth A. Integrity and disrespect: Principles of a conception of morality based on the theory of recognition. Polit Theory. 1992 May;20(2):187–201.

79. Koehler G. Transformative social protection: reflections on South Asian policy experiences. IDS Bulletin. 2011 Nov;42(6):96–103.

80. Key Indicators Database [Internet]. [place unknown]: Asian Development Bank; 2023 [cited 2023 Nov 13]. Available from: https://kidb.adb.org/

81. Parnini S. Poverty and governance in South Asia. New York (NY): Routledge; 2014 Nov 20.

82. Bynner C, Terje A. Knowledge mobilisation in public service reform: integrating empirical, technical and practical wisdom. Evid Policy. 2021;17(1):75–91.

83. Global Affairs Canada. Resilient health systems for all - concept notes [Internet]. Canada: Government of Canada; 2023 Jun 29 [cited 2023 Nov 7]. Available from: https://www.international.gc.ca/world-monde/funding-financement/resilient_health-systemes_sante.aspx?lang=eng

84. Pearse W. The problems with development aid [Internet]. [place unknown]: Inomics; 2021 Apr 7 [cited 2023 Nov 7]. Available from: https://inomics.com/blog/the-problems-with-development-aid-1388062

85. Sridhar D. Seven challenges in international development assistance for health and ways forward. J Law Med Ethics. 2010;38(3):459–69.

86. Queirós A, Faria D, Almeida F. Strengths and limitations of qualitative and quantitative research methods. Eur J Educ Stud. 2017 Sep 7;3(9):369–87.

87. Rao J. The caste system: effects on poverty in India, Nepal and Sri Lanka. Global Majority E-Journal. 2010 Dec;1(2):97–106.

88. Tamim T, Tariq H. The intersection of caste, social exclusion and educational opportunity in rural Punjab. Int J Educ Dev. 2015 Jul 1;43:51–62.

89. Imai KS, Sato T. Decentralization, democracy and allocation of poverty alleviation programmes in rural India. Eur J Dev Res. 2012 Feb 1;24(1):125–43.

90. Jha R, Bhattacharyya S, Gaiha R. Temporal variation of capture of anti-poverty programs: rural public works and food for work programs in rural India. Int Rev Appl Econ. 2011 May 1;25(3):349–62.

91. Murgai R, Zaidi S. Effectiveness of food assistance programs in Bangladesh. J Dev Soc. 2005 Jun;21(1-2):121–42.

92. Nandi A, Ashok A, Laxminarayan R. The socioeconomic and institutional determinants of participation in India’s health insurance scheme for the poor. PLoS One. 2013 Jun 21;8(6):e66296.

93. Ahmed AU, Del Ninno C. The food for education program in Bangladesh: An evaluation of its impact on educational attainment and food security. Washington (DC): International Food Policy Research Institute; 2002 Sep. 85 p.

94. National Social Assistance Programme. About us: enhancement of pension amount under IGNOAPS in 2011 [Internet]. India: Ministry of Rural Development; [cited 2023 Sep 26]. Available from: https://nsap.nic.in/circular.do?method=aboutus

95. Cheema I, Farhat M, Binci M, Asmat R, Javeed S, O’Leary S. Benazir Income Support Programme: evaluation report [Internet]. Pakistan: Government of Pakistan; 2020 [cited 2023 April 17]. Available from: https://bisp.gov.pk/SiteImage/Misc/files/BISP_EvaluationReport_Ver%20without_FINAL.pdf

96. Naqvi SM, Sabir HM, Shamim A, Tariq M. Social safety nets and poverty in Pakistan (a case study of BISP in Tehsil Mankera district Bhakkar). J Financ Econ. 2014;2(2):44–9.

97. Saeed S, Iqbal J. Challenges and issues in design and implementation: ERRA’s Livelihood Support Cash Grant Programme for vulnerable communities. Pakistan: Earthquake Reconstruction & Rehabilitation Authority; 2010. 16 p.

